# Genome wide association study based on clustering by obesity-related variables shed light on a genetic architecture of obesity in Japanese and UK population

**DOI:** 10.1101/2023.03.06.23286876

**Authors:** Ippei Takahashi, Hisashi Ohseto, Fumihiko Ueno, Tomomi Onuma, Akira Narita, Taku Obara, Mami Ishikuro, Keiko Murakami, Aoi Noda, Atsushi Hozawa, Junichi Sugawara, Gen Tamiya, Shinichi Kuriyama

**Affiliations:** Graduate School of Medicine, Tohoku University, Sendai, Japan; Tohoku Medical Megabank Organization, Tohoku University, Sendai, Japan; Tohoku University Hospital, Sendai, Japan; RIKEN Center for Advanced Intelligence Project, Tokyo, Japan; International Research Institute of Disaster Science, Tohoku University, Sendai, Japan

**Keywords:** GWAS, obesity, BMI, cGWAS, cluster analysis

## Abstract

**Background:** Many loci associated with obesity have been reported in previous genome-wide association studies (GWASs). However, it remains unclear whether variants at all these loci contributed to onset of obesity or whether one or a few variants cause obesity when obesity is a genetically heterogeneous population.

**Objective:** To investigate the genetic architecture of obesity by clustering a population with obesity into clusters using obesity-related factors.

**Methods:** This study was based on the Tohoku Medical Megabank Project Birth and Three-Generation Cohort Study and the Community-Based Cohort Study. As the Step-1, a GWAS with body mass index (BMI) as an outcome was performed for all 48,365 eligible participants. As the Step-2, we then assigned the 13,067/48,365 participants with obesity (BMI ≥ 25 kg/m^2^) using the k-prototype to 5 clusters. Obesity-related factors (such as age, nutrient intake, physical activity, sleep duration, difference between weight at age 20 and current weight, smoking, alcohol drinking, psychological distress, and birth weight) were used for clustering. Subsequently, participants in each cluster and those with a BMI < 25 kg/m^2^ were combined, and GWASs were performed according to the 5 clusters. Additionally, a sub-analysis using data from the UK Biobank was conducted to compare the results.

**Results:** The Step-1 detected 18 genes, most of which were reportedly associated with obesity or obesity-related topics in previous studies. The result of Step-2, of the 18 genes detected in Step-1, *LINC01741, CRYZL2P-SEC16B*, and *SEC16B* were significantly related to Cluster 2, *FTO, PMAIP1*, and *MC4R* to Cluster 3, and *BDNF, BDNF-AS, LINC00678*, and *KIF18A* to Clusters 4 and 5. In the sub-analysis, a similar phenomenon was observed in which separate obesity-related genes were detected for each cluster.

**Conclusions:** Our data support the notion that a decreased sample size with increased homogeneity may reveal insights into the genetic architecture of obesity.

## Introduction

Obesity is a serious global medical and economic issue that represents a major risk factor for many lifestyle-related diseases, such as diabetes, hyperlipidemia, and hypertension (1,2). The global proportion of individuals with a body-mass index (BMI) ≥ 25 kg/m^2^ is reportedly 36.9% for men and 38.0% for women (3). The pathogenesis of obesity is complex and includes regulation of calorie utilization, appetite, and physical activity, as well as health care availability, socioeconomic status, and underlying genetic and environmental factors (4,5).

The heritability of BMI has been widely reported. For instance, in twin studies, the BMI heritability ranged from 30% to 90% (6–8), whereas in genome-wide association studies (GWASs) it was estimated to be 20–30% (9–11), and only ∼3% has been elucidated based on genome wide significant loci (9,10). Although GWASs using BMI as an outcome have identified over 100 associated loci (9–14), it remains unclear whether they all contribute to the development of obesity via the same pathway. Indeed, the association of these genetic variants with obesity may be explained by a polygenic model in which the effects of each variant are weak yet contribute to the onset of obesity (15). Hence, if the genetic architecture of obesity can be explained by a polygenic model, we would expect that larger sample sizes correspond to more identified signals, whereas as fewer signals would be associated with smaller sample sizes (Supplementary Figure 1). Meanwhile, within a genetically heterogeneous population of obesity, if few variants exhibit a relatively strong influence leading to obesity in a portion of the subtypes included therein, then dividing the population with obesity into homogeneous groups could detect unique genes in each population, even with a reduced sample size. However, to our knowledge, no GWASs have been conducted by dividing persons with obesity into more homogeneous populations.

Traylor et al. demonstrated that attempts to categorize patients with a complex disease into more homogeneous subgroups provided more power to elucidate hidden heritability in a simulation study (16). Thus, clustering algorithms for machine learning could reveal novel and more genetically homogeneous clusters. Accordingly, the purpose of this study was to investigate the genetic architecture of obesity by dividing individuals with obesity into clusters using various obesity-related factors and machine learning techniques and performing GWAS on each cluster (cluster-based GWAS: cGWAS) (17,18).

## Methods

### Population

This study was conducted according to the guidelines of the Declaration of Helsinki (19), and the protocol was reviewed and approved by the Institutional Review Board of the Tohoku Medical Megabank Organization. In the main study, we used data from cohort studies conducted by the Tohoku Medical Megabank Project (TMM) Birth and Three-Generation Cohort Study (BirThree Cohort Study) and the TMM Community-Based Cohort Study (CommCohort Study) (20-22). The BirThree Cohort Study and CommCohort Study were conducted in Miyagi and Iwate Prefectures, Japan. Details of the BirThree Cohort Study and the CommCohort Study have been described elsewhere (21,22). In brief, the BirThree Cohort Study is a birth and three-generation cohort study. Pregnant women were registered between July 2013 and March 2017 (21). Additionally, pregnant women’s partners (fetus’ father), pregnant women’s parents and partner’s parents (fetus’ grandparents), as well as the fetus’ siblings and their relatives, were recruited (21). Among the BirThree Cohort Study participants, fetus’ mothers (n = 22,493), fetus’ father (n = 8,823), and fetus’ grandparents (n = 8,058) were included in this study. The TMM CommCohort study is a community-based prospective cohort study including men and women aged >20 years living in the Miyagi Prefecture, northeastern Japan (22). The type 1 survey (n = 41,097) which performed in specific municipal health check-up sites, Type 2 survey (n = 13,855) which performed in assessment centers. (22).

Participants’ data were excluded based on the following criteria: withdrew consent, failed to return the self-reported questionnaire, BMI < 18.5 kg/m^2^, missing information on the food frequency questionnaire (FFQ), extreme energy intake (energy intake > mean ± 3 standard deviation [SD]), and duplicate participation in the both BirThree Cohort Study and CommCohort Study Type-1 (the data of earlier date of participation were included). Data from eligible participants of the BirThree Cohort Study (*n* = 23,479), CommCohort Study Type-1 (*n* = 34,187), and CommCohort Study Type-2 (*n* = 12,485) were combined (*n* = 70,151). In the sub-analysis, a similar analysis was performed using the UK Biobank (UKB) data (23-25) to compare the results with those of the main study. Methods for analyzing the UKB data are described in the supplementary information.

### Genotyping, imputation, and quality control

Cohort participants were genotyped using the Affymetrix Axiom Japonica Array (v2) in 19 batches, with 50 plates set for each batch. Details pertaining to the genotyping performed in TMM have been described previously (26). Following batch genotyping, samples with a call rate < 0.95 or samples with unusually high IBD values compared to other samples, were excluded. In addition, variants with Hardy-Weinberg equilibrium (HWE) *P*-values < 1.00 × 10^−5^, minor allele frequency (MAF) < 0.01, or missing fraction > 0.01 were excluded from each batch. A direct genotype dataset in PLINK BED format was obtained by merging the genotype datasets for the 19 batches. A total of 21,541 participants with missing direct genotype data were excluded. Principal component analysis (PCA) was performed using the - -pca approx option in PLINK 2.0 (27) on the direct genotype dataset and an additional 245 participants with > 4 SD for principal components 1 or 2 were excluded. Finally, a total of 48,365 participants (BirThree Cohort Study: *n* = 11,674, CommCohort Study Type-1: *n* = 27,745, CommCohort Study Type-2: *n* = 8,946) were included in the analysis (Figure 1). Plot of participants (n = 48,365) according to principal component 1 and 2 by principal component analysis was shown Supplementary Figure 2.

**Figure 1.**
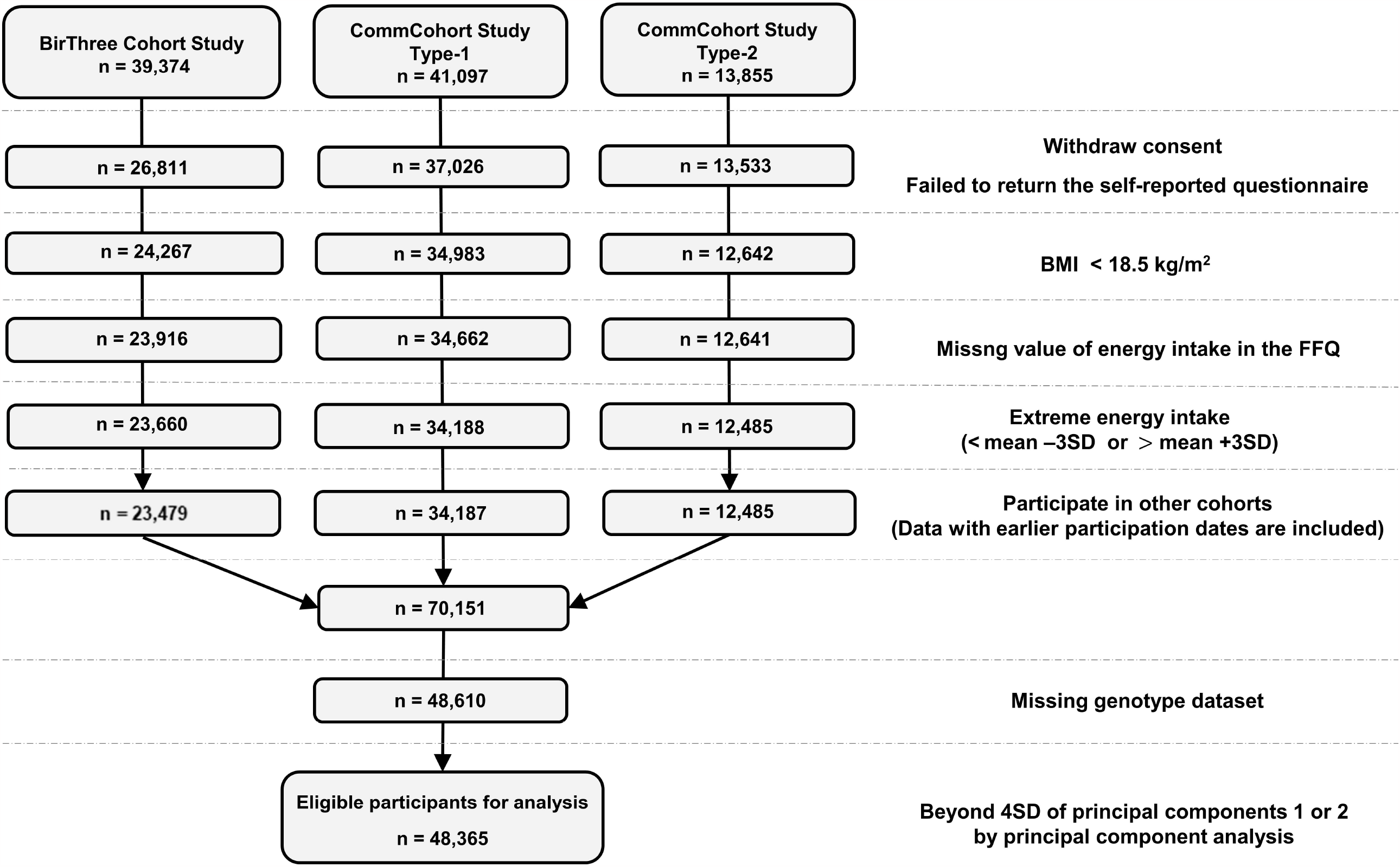
Flow chart of exclusion criteria in this study. The participants data from each cohort were excluded based on the following criteria.

To prepare an imputed genotype dataset, pre-phasing was performed using SHAPEIT2 (28), along with the --duohmm option (29), which incorporates information on the relatedness between individuals to increase phasing accuracy. The phased genotypes were subsequently imputed with a cross-imputed panel of 3.5KJPNv2 (30) and 1KGP3 (31) using IMPUTE4 (25). To create the cross-imputation panel for 3.5KJPNv2 (30) and 1KGP3 (31), the -merge_ref_panels option in IMPUTE2 was applied (32). Consequently, we obtained an imputed genotype dataset in the Oxford BGEN format (https://www.well.ox.ac.uk/gav/qctool/). For genotype imputation data, those with minor allele frequencies < 0.01 and imputation information scores < 0.8 were excluded. Finally, 9,868,333 SNPs were included in the GWASs.

### Variables

The following variables related to obesity were collected from questionnaires responded by the participants at baseline for each cohort and used for clustering: age, nutrient intake calculated from the FFQ based on frequency of food intake over the past year (energy, protein, fat, carbohydrate, sodium, potassium, calcium, magnesium, phosphorus, iron, zinc, copper, manganese, retinol equivalents, vitamin D, vitamin K, vitamin B1, vitamin B2, niacin, vitamin B6, vitamin B12, folate, pantothenic acid, vitamin C, cholesterol, dietary fiber, lycopene, α-carotene, β-carotene, and β-cryptoxanthin), frequency of leisure time physical activity (slow walking, fast walking, moderate exercise, strenuous exercise; the choices included: no activity, < once per month, 1–3 times per month, 1–2 times per week, 3–4 times per week, almost every day), time typically spent in physical activity per day (strenuous work, walk, standing, sitting) according to predetermined options (no time, min < 30, 30 ≤ min< 60, 1 ≤ h < 3, 3 ≤ h < 5, 5 ≤ h < 7, 7 ≤ h < 9, 9 ≤ h < 11, h ≥11), sleep duration (< 5 h, 5 ≤ h < 6, 6 ≤ h < 7, 7 ≤ h < 8, 8 ≤ h < 9, h ≥ 9), difference between weight at age 20 and current weight, smoking (smoked > 100 cigarettes since birth; yes or no), alcohol consumption (> 1 drink per month, quit, rarely, unable to drink), psychological distress over the past month (total K6 score [Japanese version]) (33,34), and birth weight (unknown, 1500 ≤ g < 2000, 2000 ≤ g < 2500, 2500 ≤ g< 3000, 3000 ≤ g < 3500, 3500 ≤ g < 4000, g ≥ 4000). In addition, cohort type (BirThree Cohort Study, CommCohort study Type-1 and CommCohort study Type-2) was added to the variables for the clustering.

The missing variables used for clustering were imputed using the k-nearest neighbor (KNN) algorithm (35). KNN selects k samples close to the missing values in the feature space and imputes the median of the k samples in the case of continuous variables, or the most frequent category among the k samples in the case of categorical variables. KNN was implemented using the R package VIM (36). Based on previous reports (35,37), we set k to 219 as an odd number close to the square root of 48,365 participants.

### Body mass index

BMI was computed by dividing weight (kg) by the squared height (m^2^) using self-reported height and weight on a questionnaire responded by the participants at baseline for each cohort. A BMI > 25 kg/m^2^ was defined as obese based on the Western Pacific Region of the World Health Organization criteria for Asians (38).

### Cluster analysis

The k-prototype is a clustering algorithm that combines k-means and k-modes and enables clustering using continuous and categorical variables (39). The k-prototype was implemented using the R package clustMixType (40). The number of clusters was set to 5. Continuous variables were standardized by subtracting the mean of each variable and dividing it by the SD before clustering.

### Genome-wide association study

The GWASs with BMI as a continuous variable were conducted in 2 steps. Step-1: GWAS was performed on all 48,365 participants. Step-2: 13,067 of the 48,365 participants were clustered using the k-prototype to 5 clusters. Thereafter, participants in each of the 5 obesity clusters and those with a BMI < 25 kg/m^2^ were combined, and the GWASs were performed for each of the 5 clusters (Supplementary Figure 3). To identify associations between autosomal SNPs and BMI, fastGWA with the GCTA software were employed (41).

FastGWA is a linear mixed model using a sparse genetic relationship matrix (GRM) that is reportedly robust for population stratification and familial relationships (42). The top 20 principal components calculated from the PCA of the direct genotyping dataset, sex, age, and cohort type (BirThree Cohort Study, CommCohort study type-1 and CommCohort study type-2) were included as covariates. We set the Bonferroni genome-wide significance line at *P* < 8.33×10^−9^ (5.0 × 10^−8^/6) as six GWASs were performed for Step-1 and Step-2. The detected SNPs were annotated using the ANNOVAR (43). Manhattan plots and quantile-quantile plots (Q-Q plots) were generated using R (version 4.1.0).

### Results Clustering

Following assignment of the 13,067 participants with obesity into 5 clusters, Cluster 1 contained 628 participants, Cluster 2 had 3,073, Cluster 3 had 4,111, Cluster 4 had 2,468, and Cluster 5 contained 2,787. Table 1 shows the characteristics of obese participants (BMI ≥ 25 kg/m^2^) in each cluster. The variables were characterized by mean and SD for continuous variables and by number and percentage for categorical variables. The participants in Cluster 1 had the highest energy and nutrient intakes, as well as a higher frequency of leisure-time exercise. Cluster 2 was characterized by a higher proportion of women, older age, and the highest percentage of nonsmokers. Cluster 3 participants had the lowest energy and nutrient intake and a high proportion who did not exercise during leisure time nor perform their usual physical activities (strenuous work, walking, and standing). Cluster 4 had the second-highest energy and nutrient intake. Cluster 5 was characterized by the largest proportion of men, lowest age, longest time spent standing or sitting, highest number of smokers, highest proportion of alcohol drinkers, and highest scores for psychological distress.

**Table 1.**
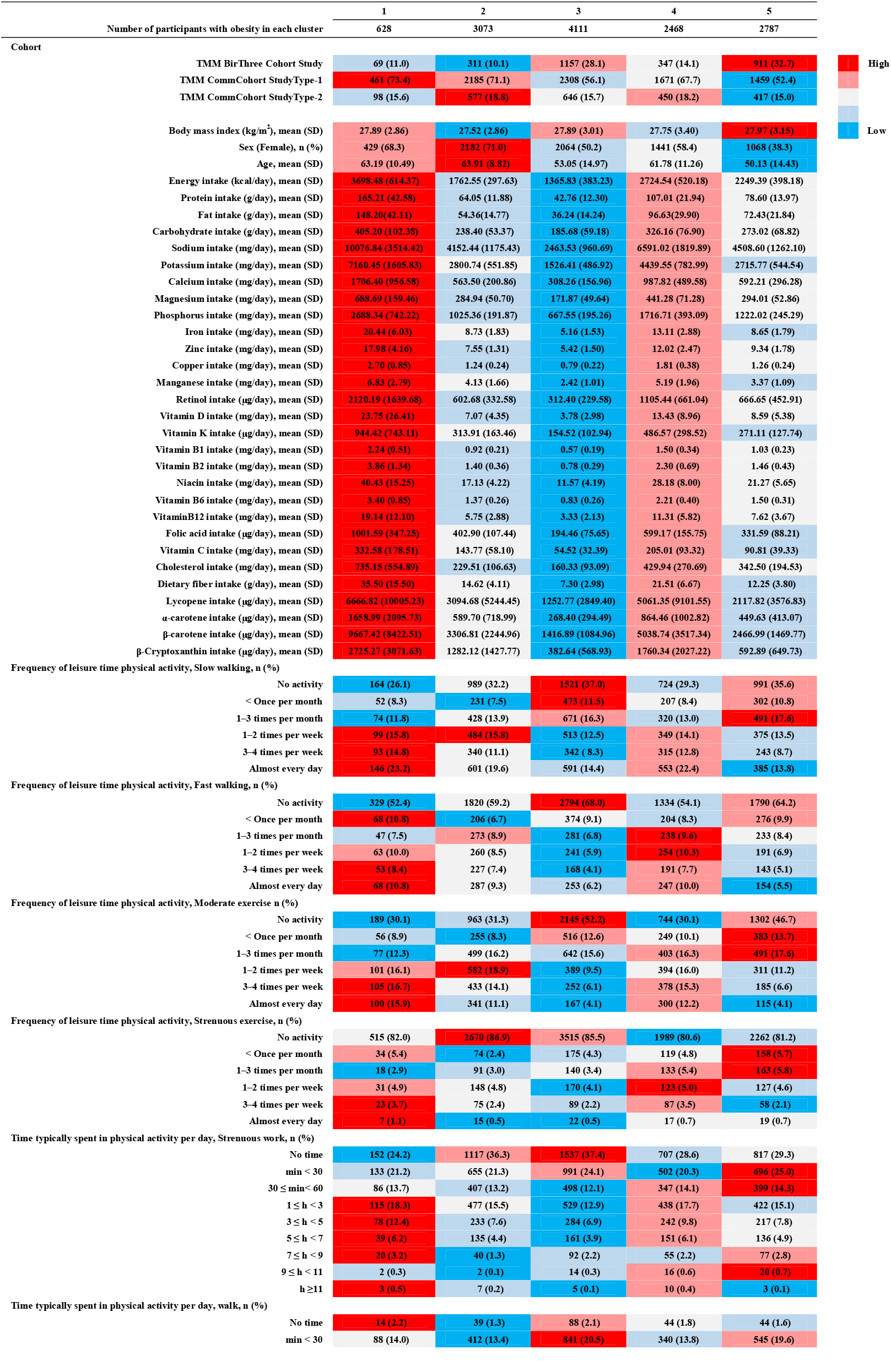
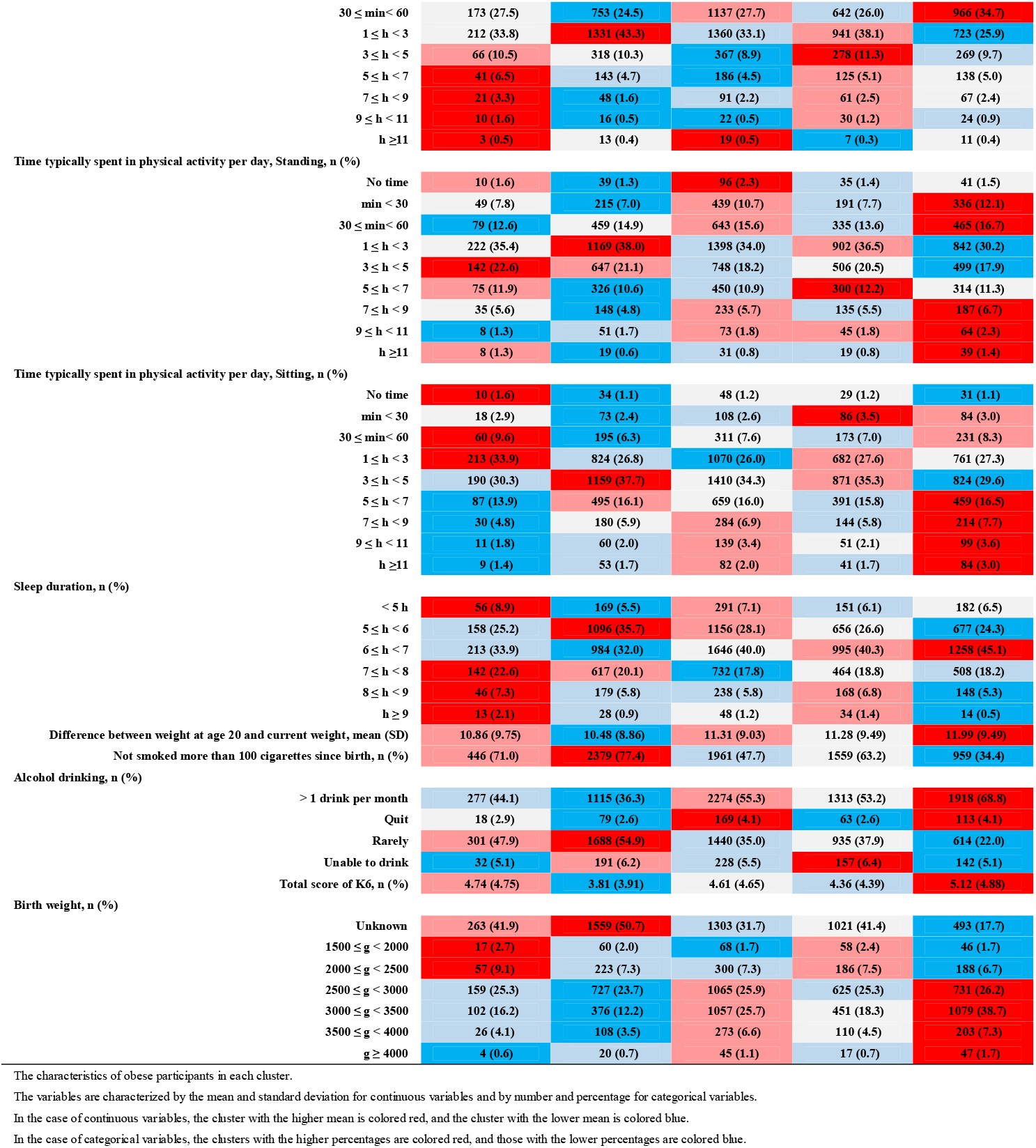
Characteristics of the clusters.

### Gene interpretation

We observed several genes that satisfied the *P* < 8.33 × 10^−9^ threshold in Step-1 (Figure 2 and Supplementary Table 1). Most genes for which associations were detected in Step-1 are reportedly associated with obesity. More specifically, *LINC01741* (44–46) (Chr 1), *CRYZL2P-SEC16B* (45,47) (Chr 1), *SEC16B* (46,47) (Chr 1), *TMEM18* (48,49) (Chr 2), *BDNF* (45,47) (Chr 11), *LINC00678* (50,51) (Chr 11), *BDNF-AS* (45,47) (Chr 11), *FTO* (9,10,45,52) (Chr 16), *MC4R* (53,54) (Chr 18), *GIPR* (13) (Chr 19), and *FBXO46* (50) (Chr 19) were previously associated with BMI. Meanwhile, *KIF18A* (Chr 11) was previously associated with visceral fat (55), *PMAIP1* (Chr 18) with serum IgE measurement (56) and monocyte count (57,58), *RSPH6A* (Chr 19) with high and low density lipoprotein cholesterol levels (59), *SYMPK* (Chr 19) with Type 2 diabetes mellitus (60) and total cholesterol levels (50), and *FOXA3* (Chr 19) with waist-to-hip ratio adjusted for BMI (52).

**Figure 2.**
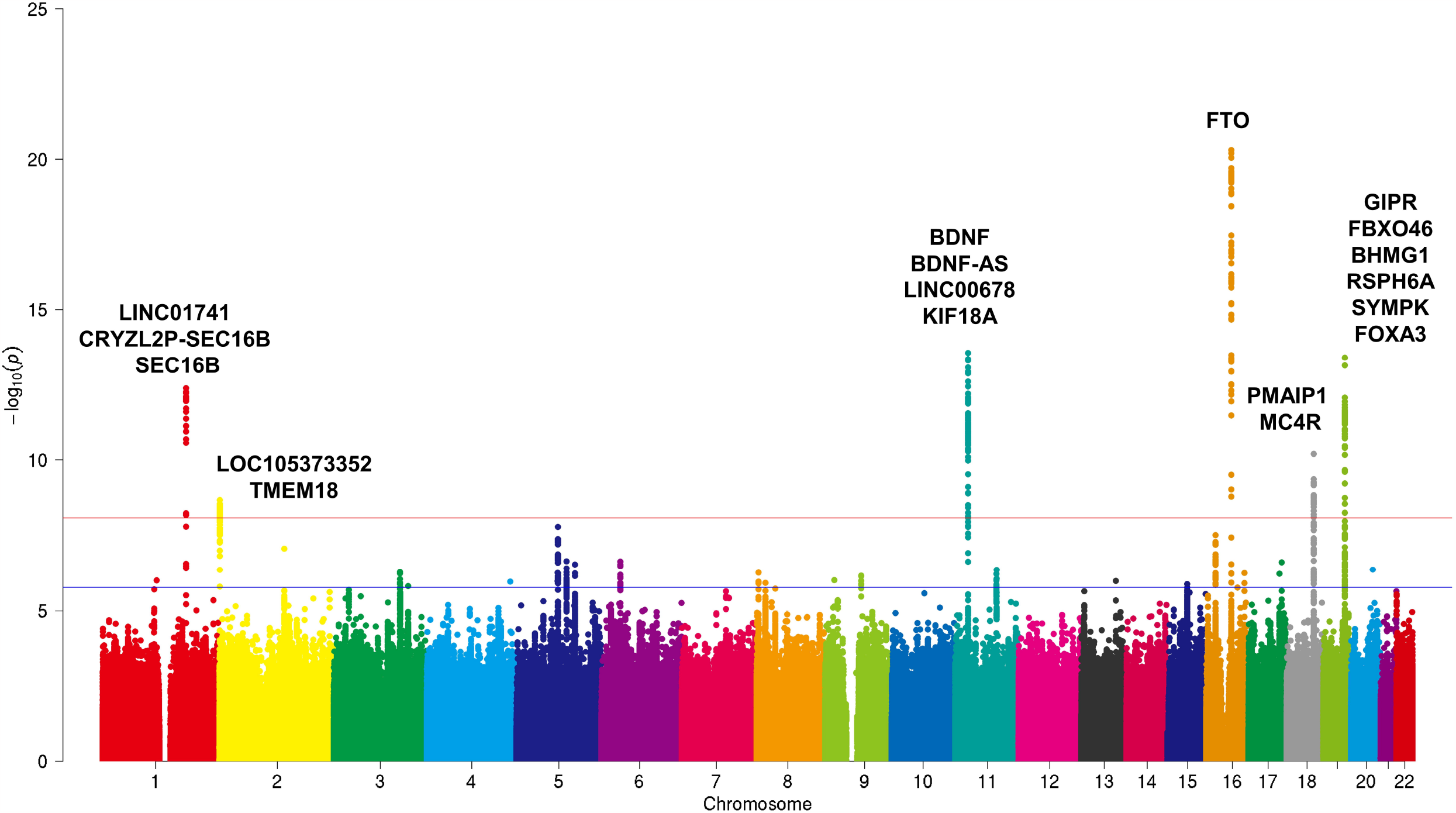
Manhattan plot of Step-1. A GWAS with BMI as a continuous variable was performed on 48,365 participants.

From the GWAS results in Step-2, several variants detected in Step-1 were observed in separate clusters (Figure 3, Supplementary Table 2). Genome-wide associations were not detected in Cluster 1. In Cluster 2, the loci that satisfied this threshold were identified as *LINC01741, CRYZL2P-SEC16B* (Chr 1; intergenic), *CRYZL2P-SEC16B*, and *SEC16B* (Chr 1). Meanwhile, in Cluster 3, the *FTO* (chromosome 16), *PMAIP1*, and *MC4R* (chromosome 18; intergenic) loci were identified. For Cluster 4, *BDNF* (Chr 11), *BDNF-AS* (Chr 11), *BDNF-AS, LINC00678* (Chr 11), and *BDNF, KIF18A* (Chr 11) loci were identified. Additionally, in Cluster 5, *BDNF-AS, LINC00678* (Chr 11), *LINC00678* (Chr 11), *BDNF-AS* (Chr 11), *BDNF* (Chr 11), and *BDNF, KIF18A* (Chr 11) loci were identified (Figure 3). Quantile-quantile plots corresponding to the GWAS results of the main study are shown in Supplementary Figure 4.

**Figure 3.**
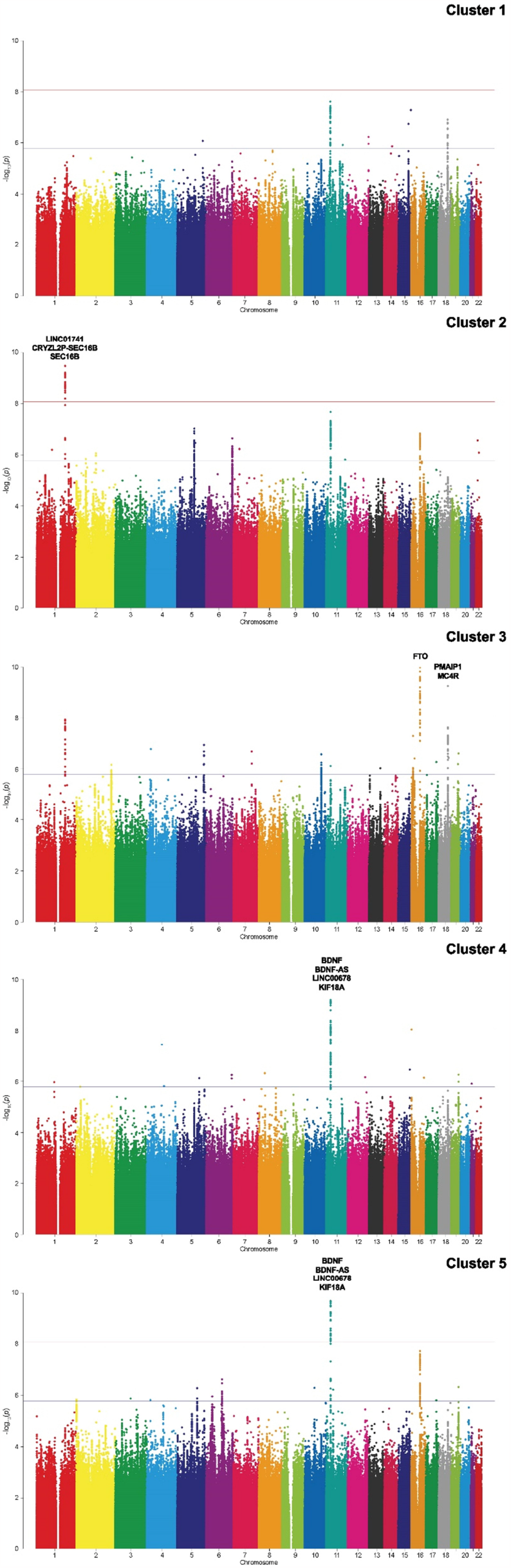
Manhattan plots of Step-2. We clustered 13,067 of the 48,356, individuals with BMI≥25kg/m^2^ using the k-prototype. Thereafter, participants with obesity in each of the 5 clusters and those with BMI< 25kg/m^2^ were then combined and GWAS was performed according to the 5 clusters (cluster-based GWAS: cGWAS).

In the sub-analysis, the UKB data was applied for comparison with the main study results. In Step-1, we confirmed the association between representative obesity-related genes and BMI (Supplementary Table 3 and Supplementary Figure 5). In Step-2, the clustering results for the 32,779 obese participants revealed that revealed that Clusters 1–5 comprised 5,874, 6,497, 6,919, 6,733, and 6,756 participants, respectively. The characteristics of each cluster are shown in Supplementary Table 4. In the GWAS results for Step-2, several variants detected in Step-1 were found in separate clusters, similar to the TMM cohort analysis (Supplementary Table 5 and Supplementary Figure 6).

## Discussion

Herein, we conducted a GWAS of all participants for BMI in Step-1. In Step-2, obese participants (BMI ≥ 25 kg/m^2^) were divided into 5 clusters based on obesity-related-factors, and GWAS was performed for each of the 5 clusters. Consequently, several genes identified in previous studies were confirmed in Step-1. Of the 18 genes detected in Step-1, *LINC01741, CRYZL2P-SEC16B*, and *SEC16B* were significantly associated with Cluster 2, *FTO, PMAIP1*, and *MC4R* to Cluster 3, and *BDNF, BDNF-AS, BDNF-AS, LINC00678*, and *KIF18A* to Clusters 4 and 5. A similar phenomenon was observed in the sub-analysis using UKB data, in which unique obesity-related genes were detected in each cluster.

It is important to consider how the cluster characteristics relate to the variants identified in each cluster. Indeed, the GWAS results in Step-2 may be partially explained by cluster characteristics. In cluster 1, significant associations were not detected, which might be due to the low number of participants with a BMI > 25.0 kg/m^2^ as this cluster contained the fewest obese participants. Hence, the detection power would have been insufficient.

*FTO, PMAIP1*, and *MC4R* (intergenic) variants were associated with BMI in Cluster Variants in the *FTO* region regulate *IRX3* and *IRX5* expression (61), which promotes fat accumulation and cause obesity. Meanwhile, melanocortin-4-receptors (MC4R) transcribed by the *MR4C* gene regulate food intake and energy expenditure (62,63). Moreover, *MC4R* in the paraventricular hypothalamus or amygdala controls food intake, while its expression elsewhere is responsible for energy expenditure (62). Therefore, the genetic variants in *FTO* and *MC4R*, which have been attributed to increased body fat accumulation and reduced energy expenditure, may partially account for the obesity of individuals in Cluster 3 despite a low energy intake.

In Cluster 4 and Cluster 5, *BDNF* and *BDNF-AS* variants were identified. Obese participants in Cluster 4 had the second highest energy and nutrient intake, while those in Cluster 5 had the highest mean score for psychological distress (K6 total score). Brain derived neurotrophic factor (BDNF), which is transcribed by the *BDNF* gene, promotes the development and growth of nerve cells and reportedly has anti-obesity effects (64,65). Furthermore, transcription of the *BDNF-AS* (antisense RNA) gene is responsible for regulating *BDNF* expression (66). Thus, altering BDNF regulation might affect the central nervous system and alter eating behaviors and psychiatric conditions, as seen in this cluster.

In Cluster 2, *SEC16B* variants were detected; the obese participants in this cluster had the highest proportion of women, increased age, and nonsmokers. Variants of *SEC16B* may be associated with obesity via regulation of dietary lipid absorption and appetite (67,68). However, to our knowledge, no previous data has made direct connections between the characteristics of Cluster 2 participants and SEC16B variants. On the other hand, it should be noted that the characteristics of clusters are not always recognizable to humans. That is, given that clustering algorithms extract latent features by combining numerous variables, the resulting clusters, although more homogeneous, are not necessarily comprehensible. Thus, it will be necessary to define these obscure clusters identified by clustering algorithms.

This study has several strengths. First, the GWAS results had high validity. That is, most genes detected in this study were previously reported to be associated with BMI. Therefore, the GWAS data was considered appropriate. Second, the TMM and UKB cohorts had various obesity-related factors. Using these 2 cohorts, it was possible to cluster the obese population into more homogeneous groups using a rich set of obesity-related factors. Third, the sub-analysis replicated the phenomenon, in which unique obesity-related genes were detected in each cluster. This supports the hypothesis that obesity is an aggregation of heterogeneous subgroups. The findings of this study suggest possibility that by dividing obesity into homogeneous populations, fewer genetic variants could explain obesity in each subgroup. Although many issues remain to be addressed to elucidate the full genetic architecture of obesity, the current study provides important insights with the potential to inform the development of personalized treatment or nutritional support for obesity. More specifically, once clusters are identified, a classifier can be created using the cluster numbers as training data, which can then be applied to classify obesity into subgroups and verify the effectiveness of obesity treatment according to the subgroups.

## Limitations

This study has certain limitations. First, it is unclear whether the selection of variables, algorithm, or number of clusters was optimal. In this study, many factors related to obesity were selected, however, the existence of unknown obesity-related factors cannot be ruled out. In addition, the number of clusters in this study was arbitrarily set to 5. Therefore, it is needed to explore them in the future. Second, obesity was assessed at a temporal point; therefore, the possibility of misclassification may have occurred. Even those who were not obese at the time of measurement had the potential to become obese with age. Third, the BMI was calculated using height and weight from self-reported questionnaires in the main study. Previous studies have shown no substantial differences between BMI calculated from self-reported height and weight and that calculated from measured height and weight, indicating that self-reported weight and height are useful (69). Therefore, it is unlikely that the use of self-reported height and weight data significantly distorted the results. Fourth, we could not assess the heritability of each cluster due to the small sample size.

## Conclusion

Our data suggest that a decreased sample size with increased homogeneity may reveal insights into the genetic architecture of obesity.

## Supporting information

Supplementary Figure 1. Virtual Manhattan plots with different number of participants.

Supplementary Figure 2. Plot of participants according to principal component 1 and 2 by principal component analysis.

Supplementary Figure 3. Details of the cluster-based GWAS.

Supplementary Figure 4. quantile-quantile plots and lambda values in main study.

Supplementary Figure 5. Manhattan plots (a) and corresponding quantile-quantile plots (b) of Step-1 in sub-analysis.

Supplementary Figure 6. Manhattan plots (a) and corresponding quantile-quantile plots (b) of Step-2 in sub-analysis.

Supplementary documentation

## Data Availability

For the TMM biobank, data are available from the authors upon reasonable request and with the permission of the TMM biobank. All inquiries about access to the data should be sent to the TMM biobank. For the UKB, the data will be available to the public by requesting it from UK Biobank.

## Abbreviations

AS: antisense RNA
BDNF: brain-derived neurotrophic factor
BirThree Cohort Study: Birth and Three-Generation Cohort Study, CommCohort Study
BMI: body mass index
cGWAS: cluster-based GWAS
FFQ: food frequency questionnaire
GRM: genetic relationship matrix
GWAS: genome-wide association study
HWE: Hardy-Weinberg equilibrium
KNN: k-nearest neighbors
MAF: minor allele frequency
MC4R: melanocortin-4-receptors
PCA: principal component analysis
SD: standard deviation
TMM: Tohoku Medical Megabank Project, Community-Based Cohort Study

## Acknowledgments

The authors thank the participants of the Tohoku Medical Megabank Project Birth and Three-Generation Cohort Study, Community-Based Cohort Study, and UK Biobank. The authors also thank the staff members of the Tohoku Medical Megabank Organization (https://www.megabank.tohoku.ac.jp/english/a210901/) and UK Biobank.

## Author Contributions

IT, HO, FU, TO, and SK designed research; IT conducted research; IT analyzed data; and IT HO, and SK wrote the paper. SK had primary responsibility for final content. All authors read and approved the final manuscript.

## Figure Legends

**Supplementary Figure 1. Virtual Manhattan plots with different number of participants**.

**Supplementary Figure 2. Plot of participants according to principal component 1 and 2 by principal component analysis**.

**Supplementary Figure 3. Details of the cluster-based GWAS**.

**Supplementary Figure 4. quantile-quantile plots and lambda values in main study**.

**Supplementary Figure 5. Manhattan plots (a) and corresponding quantile-quantile plots**

**(b) of Step-1 in sub-analysis**.

**Supplementary Figure 6. Manhattan plots (a) and corresponding quantile-quantile plots (b) of Step-2 in sub-analysis**.

## References

1. Haslam DW, James WPT. Obesity. Lancet. 2005;366:1197–209. doi:10.1016/S0140-6736(05)67483-1

2. Eckel RH, Grundy SM, Zimmet PZ. The metabolic syndrome. Lancet. 2005;365:1415–28. doi:10.1016/S0140-6736(05)66378-7

3. Ng, M, Fleming T, Robinson M, Thomson B, Graetz N, Margono EC, et al. Global, regional, and national prevalence of overweight and obesity in children and adults during 1980-2013: a systematic analysis for the Global Burden of Disease Study 2013. Lancet. 2014;384:766–81. doi:10.1016/S0140-6736(14)60460-8

4. Lin X, Li H. Obesity: Epidemiology, pathophysiology, and therapeutics. Front Endocrinol (Lausanne). 2021;12:706978. doi:10.3389/fendo.2021.706978

5. Lyon HN, Hirschhorn JN. Genetics of common forms of obesity: a brief overview. Am J Clin Nutr. 2015;82:215S–7S. doi:10.1093/ajcn/82.1.215S

6. Feng R. How much do we know about the heritability of BMI?. Am J Clin Nutr. 2016;104:243–4. doi:10.3945/ajcn.116.139451

7. Elks CE, den Hoed M, Zhao JH, Sharp SJ, Wareham NJ, Loos RJF, et al. Variability in the heritability of body mass index: a systematic review and meta-regression. Front Endocrinol (Lausanne). 2012;3:29. doi:10.3389/fendo.2012.00029

8. Min J, Chiu DT, Wang Y. Variation in the heritability of body mass index based on diverse twin studies: a systematic review. Obes Rev 2013;14:871–82. doi:10.1111/obr.12065

9. Akiyama M, Okada Y, Kanai M, Takahashi A, Momozawa Y, Ikeda M, et al. Genome-wide association study identifies 112 new loci for body mass index in the Japanese population. Nat Genet. 2017;49:1458–6. doi:10.1038/ng.3951

10. Locke AE, Kahali B, Berndt SI, Justice AE, Pers TH, Day FR, et al. Genetic studies of body mass index yield new insights for obesity biology. Nature 2015;518:197–206. doi:10.1038/nature14177.

11. Yang J, Bakshi A, Zhu Z, Hemani G, Vinkhuyzen AE, Lee SH, et al. Genetic variance estimation with imputed variants finds negligible missing heritability for human height and body mass index. Nat Genet. 2015;47:1114–20. doi:10.1038/ng.3390

12. Speliotes EK, Willer CJ, Berndt SI, Monda KL, Thorleifsson G, Jackson AU, et al. Association analyses of 249,796 individuals reveal 18 new loci associated with body mass index. Nat Genet. 2010;42:937–48. doi:10.1038/ng.686

13. Wen W, Zheng W, Okada Y, Takeuchi F, Tabara Y, Hwang J-Y, et al. Meta-analysis of genome-wide association studies in East Asian ancestry populations identifies four new loci for body mass index. Hum Mol Genet. 2014;23:5492–504. doi:10.1093/hmg/ddu248

14. Scuteri A, Sanna S, Chen W-M, Uda M, Albai G, Strait J, et al. Genome-wide association scan shows genetic variants in the FTO gene are associated with obesity-related traits. PLoS Genet. 2007;3:e115. doi:10.1371/journal.pgen.0030115.

15. Khera AV, Chaffin M, Wade KH, Zahid S, Brancale J, et al. Polygenic prediction of weight and obesity trajectories from birth to adulthood. Cell. 2019;177:587–96. doi:10.1016/j.cell.2019.03.028

16. Traylor M, Markus H, Lewis CM. Homogeneous case subgroups increase power in genetic association studies. Eur J Hum Genet. 2015;23:863. doi:10.1038/ejhg.2014.194

17. Ueno F, Onuma T, Takahashi I, Ohseto H, Narita A, Obara T, et al. Deep embedded clustering by relevant scales and genome-wide association study in autism. bioRxiv. 2022. doi:10.1101/2022.07.25.500917.

18. Narita A, Nagai M, Mizuno S, Ogishima S, Tamiya G, Ueki M, et al. Clustering by phenotype and genome-wide association study in autism. Transl Psychiatry. 2020;10:290. doi:10.1038/s41398-020-00951-x

19. World Medical Association. World Medical Association Declaration of Helsinki: ethical principles for medical research involving human subjects. JAMA 2013;310:2191–4. doi:10.1001/jama.2013.281053.

20. Kuriyama S, Yaegashi N, Nagami F, Arai T, Kawaguchi Y, Osumi N, et al. The Tohoku Medical Megabank Project: Design and mission. J Epidemiol. 2016;26:493–511. doi:10.2188/jea.JE20150268

21. Kuriyama S, Metoki H, Kikuya M, Obara T, Ishikuro M, Yamanaka C, et al. Cohort Profile: Tohoku Medical Megabank Project Birth and Three-Generation Cohort Study (TMM BirThree Cohort Study): rationale, progress and perspective. Int J Epidemiol. 2020;49:18–9. doi:10.1093/ije/dyz169

22. Hozawa A, Tanno K, Nakaya N, Nakamura T, Tsuchiya N, Hirata T, et al. Study Profile of the Tohoku Medical Megabank Community Based Cohort Study. J Epidemiol. 2021;31:JE20190271. doi:10.2188/jea.JE20190271

23. Sudlow C, Gallacher J, Allen N, Beral V, Burton P, Danesh J, et al. UK Biobank: an open access resource for identifying the causes of a wide range of complex diseases of middle and old age. PLoS Med. 2015;12:e1001779. doi:10.1371/journal.pmed.1001779

24. Bycroft C, Freeman C, Petkova D, Band G, Elliott LT, Sharp K, et al. Genome-wide genetic data on ∼500,000 UK Biobank participants. bioRxiv. 2017. doi:10.1101/166298.

25. Bycroft C, Freeman C, Petkova D, Band G, Elliott LT, Sharp K, et al. The UK Biobank resource with deep phenotyping and genomic data. Nature 2018;562:203–9. doi:10.1038/s41586-018-0579-z

26. Yamada M, Motoike IN, Kojima K, Fuse N, Hozawa A, Kuriyama S, et al. Genetic loci for lung function in Japanese adults with adjustment for exhaled nitric oxide levels as airway inflammation indicator. Commun Biol. 2021;15:1288. doi:10.1038/s42003-021-02813-8

27. Chang CC, Chow CC, Tellier LC, Vattikuti S, Purcell SM, Lee JJ. Second-generation PLINK: Rising to the challenge of larger and richer datasets. Gigascience. 2015;4:7. doi:10.1186/s13742-015-0047-8

28. Delaneau O, Zagury JF, Marchini J. Improved whole-chromosome phasing for disease and population genetic studies. Nat Methods. 2013;10:5–6. doi:10.1038/nmeth.2307.

29. O’Connell J, Gurdasani D, Delaneau O, Pirastu N, Ulivi S, Cocca M, et al. A general approach for haplotype phasing across the full spectrum of relatedness. PLoS Genet. 2014;10:e1004234. doi:10.1371/journal.pgen.1004234.

30. Tadaka S, Katsuoka F, Ueki M, Kojima K, Makino S, Saito S, et al. 3.5KJPNv2: an allele frequency panel of 3552 Japanese individuals including the X chromosome. Hum Genome Var. 2019;6:28. doi:10.1038/s41439-019-0059-5

31. The 1000 Genomes Project Consortium. A global reference for human genetic variation. Nature. 2015;526:68–74. doi:10.1038/nature15393

32. Howie BN, Donnelly P, Marchini J. A flexible and accurate genotype imputation method for the next generation of genome-wide association studies. PLoS Genet. 2009;5:e1000529. doi:10.1371/journal.pgen.1000529

33. Kessler RC, Andrews G, Colpe LJ, Hiripi E, Mroczek DK, Normand SLT, et al. Short screening scales to monitor population prevalences and trends in non-specific psychological distress. Psychol Med. 2002;32:959–76. doi:10.1017/s0033291702006074

34. Furukawa TA, Kawakami N, Saitoh M, Ono Y, Nakane Y, Nakamura Y, et al. The performance of the Japanese version of the K6 and K10 in the World Mental Health Survey Japan. Int J Methods Psychiatr Res. 2008;17:152–8. doi:10.1002/mpr.257

35. Zhang, Z. Introduction to machine learning: k-nearest neighbors. Ann Transl Med. 2016;4:2188. doi:10.21037/atm.2016.03.37.

36. Templ M, Kowarik A, Alfons A, de Cillia G, Prantner B, Rannetbauer W. Visualization and imputation of missing values. 2022. Available from: https://cran.r-project.org/web/packages/VIM/VIM.pdf

37. Lantz B. Machine learning with R: Expert techniques for predictive modeling to solve all your data analysis problems. 2nd ed. Birmingham-Mumbai: Packt Publishing, 2015.

38. World Health Organization. Regional Office for the Western Pacific. The Asia-Pacific perspective: redefining obesity and its treatment. Sydney: Health Communications Australia, 2000. Available from: https://apps.who.int/iris/handle/10665/206936

39. Huang Z. Extensions to the k-Means Algorithm for Clustering Large Data Sets with Categorical Values. Data Min Knowl Discov. 1998;2:283–304. doi:10.1023/A:1009769707641

40. Package ‘clustMixType’ k-Prototypes Clustering for Mixed Variable-Type Data, 2021. Available from: https://cran.r-project.org/web/packages/clustMixType/clustMixType.pdf

41. Wang Y, Ding X, Tan Z, Ning C, Xing K, Yang T, et al. Genome-wide association study of piglet uniformity and farrowing interval. Front Genet. 2017;8:194. doi:10.3389/fgene.2017.00194

42. Jiang L, Zheng Z, Fang H, Yang J. A generalized linear mixed model association tool for biobank-scale data. Nat Genet. 2021;53:1616–21. doi:10.1038/s41588-021-00954-4

43. Wang K, Li M, Hakonarson H. ANNOVAR: functional annotation of genetic variants from high-throughput sequencing data. Nucleic Acids Res. 2010;38:e164 doi:10.1093/nar/gkq603

44. Ng MCY, Graff M, Lu Y, Justice AE, Mudgal P, Liu C-T, et al. Discovery and fine-mapping of adiposity loci using high density imputation of genome-wide association studies in individuals of African ancestry: African Ancestry Anthropometry Genetics Consortium. PLoS Genet. 2017;13:e1006719. doi:10.1371/journal.pgen.1006719

45. Wojcik GL, Graff M, Nishimura KK, Tao R, Haessler J, Gignoux CR, et al. Genetic analyses of diverse populations improves discovery for complex traits. Nature 2019;570:514–8. doi:10.1038/s41586-019-1310-4

46. Monda KL, Chen GK, Taylor KC, Palmer C, Edwards TL, Lange LA, et al. A meta-analysis identifies new loci associated with body mass index in individuals of African ancestry. Nat Genet. 2013;45:690–6. doi:10.1038/ng.2608

47. Thorleifsson G, Walters GB, Gudbjartsson DF, Steinthorsdottir V, Sulem P, Helgadottier A, et al. Genome-wide association yields new sequence variants at seven loci that associate with measures of obesity. Nat Genet. 2008;41:18–24. doi:10.1038/ng.274

48. Pei YF, Zhang L, Liu Y, Li J, Shen H, Liu Y-Z, et al. Meta-analysis of genome-wide association data identifies novel susceptibility loci for obesity. Hum Mol Genet. 2014;23:820–30. doi:10.1093/hmg/ddt464

49. Willer CJ, Speliotes EK, Loos RJF, Li S, Lindgren CM, Heid IM, et al. Six new loci associated with body mass index highlight a neuronal influence on body weight regulation. Nat Genet. 2008;41:25–34. doi:10.1038/ng.287

50. Zhu Z, Guo Y, Shi H, Liu C-L, Panganiban RA, Chung W, et al. Shared genetic and experimental links between obesity-related traits and asthma subtypes in UK Biobank. J Allergy Clin Immunol. 2020;145:537–49. doi:10.1016/j.jaci.2019.09.035

51. Tachmazidou I, Süveges D, Min JL, Ritchie GRS, Steinberg J, Walter K, et al. Whole-genome sequencing coupled to imputation discovers genetic signals for anthropometric traits. Am J Hum Genet. 2017;100:865–84. doi:10.1016/j.ajhg.2017.04.014

52. Sakaue S, Kanai M, Tanigawa Y, Karjalainen J, Kurki M, Koshiba S, et al. A cross-population atlas of genetic associations for 220 human phenotypes. Nat Genet. 2021;53:1415–24. doi:10.1038/s41588-021-00931-x.

53. Barton AR, Sherman MA, Mukamel RE, Loh PR. Whole-exome imputation within UK Biobank powers rare coding variant association and fine-mapping analyses. Nat Genet. 2021;53:1260–9. doi:10.1038/s41588-021-00892-1

54. Akbari P, Gilani A, Sosina O, Kosmicki JA, Khrimian L, Fang Y-Y, et al. Sequencing of 640,000 exomes identifies GPR75 variants associated with protection from obesity. Science 2021;373:eabf8683. doi:10.1126/science.abf8683.

55. Shin J, Syme C, Wang D, Richer L, Pike GB, Gaudet D, et al. Novel genetic locus of visceral fat and systemic inflammation. J Clin Endocrinol Metab. 2019;104:3735–42. doi:10.1210/jc.2018-02656

56. Akenroye AT, Brunetti T, Romero K, Daya M, Kanchan K, Shankar G, et al. Genome-wide association study of asthma, total IgE, and lung function in a cohort of Peruvian children. J Allergy Clin Immunol. 2021;148:1493–504. doi:10.1016/j.jaci.2021.02.035.

57. Chen MH, Raffield LM, Mousas A, Sakaue S, Huffman JE, Moscati A, et al. Trans-ethnic and ancestry-specific blood-cell genetics in 746,667 individuals from 5 global populations. Cell. 2020;182:1198–213. doi:10.1016/j.cell.2020.06.045

58. Vuckovic D, Bao EL, Akbari P, Lareau CA, Mousas A, Jiang T, et al. The polygenic and monogenic basis of blood traits and diseases. Cell. 2020;182:1214–31. doi:10.1016/j.cell.2020.08.008

59. Sinnott-Armstrong N, Tanigawa Y, Amar D, Mars N, Benner C, Aguirre M, et al. Genetics of 35 blood and urine biomarkers in the UK Biobank. Nat Genet. 2021;53:185–94. doi:10.1038/s41588-020-00757-z

60. Mahajan A, Taliun D, Thurner M, Robertson NR, Torres JM, Rayner NW, et al. Fine-mapping type 2 diabetes loci to single-variant resolution using high-density imputation and islet-specific epigenome maps. Nat Genet. 2018;50:1505–13 doi:10.1038/s41588-018-0241-6

61. Claussnitzer M, Dankel SN, Kim K-H, Quon G, Meuleman W, Haugen C, et al. FTO obesity variant circuitry and adipocyte browning in humans. N Engl J Med. 2015;373:895–907. doi:10.1056/NEJMoa1502214

62. Balthasar N, Dalgaard LT, Lee CE, Yu J, Funahashi H, Williams T, et al. Divergence of melanocortin pathways in the control of food intake and energy expenditure. Cell. 2005;123:493–505. doi:10.1016/j.cell.2005.08.035

63. Krashes MJ, Lowell BB, Garfield AS. Melanocortin-4 receptor–regulated energy homeostasis. Nat Neurosci. 2016;19:206–19. doi:10.1038/nn.4202

64. Noble EE, Billington CJ, Kotz CM, Wang CF. The lighter side of BDNF. Am J Physiol Regul Integr Comp Physiol. 2011;300:R1053. doi:10.1152/ajpregu.00776.2010

65. Pandit M, Behl T, Sachdeva M, Arora S. Role of brain derived neurotropic factor in obesity. Obes Med. 2020;17:100189. Doi:10.1016/j.obmed.2020.100189

66. Ghafouri-Fard S, Khoshbakht T, Taheri M, Ghanbari M. A concise review on the role of BDNF-AS in human disorders. Biomed Pharmacother. 2021;142:112051. doi:10.1016/j.biopha.2021.112051

67. Shi R, Lu W, Tian Y, Wang B, Ave L. Intestinal SEC16B modulates obesity by controlling dietary lipid absorption. bioRxiv. 2021. doi:10.1101/2021.12.07.471468.

68. Hotta K, Nakamura M, Nakamura T, Matsuo T, Nakata Y, Kamohara S, et al. Association between obesity and polymorphisms in SEC16B, TMEM18, GNPDA2, BDNF, FAIM2 and MC4R in a Japanese population. J Hum Genet. 2009;54:727–31. doi:10.1038/jhg.2009.106.

69. Haakstad LAH, Stensrud T, Gjestvang C. Does self-perception equal the truth when judging own body weight and height? Int J Environ Res Public Health. 2021;18:8502. doi:10.3390/ijerph18168502.

