## Supplementary figures and images for "Genome wide association study based on clustering by obesity-related variables shed light on a genetic architecture of obesity in Japanese and UK population"

### Supplementary Figure 1. Virtual Manhattan plots with different number of participants.

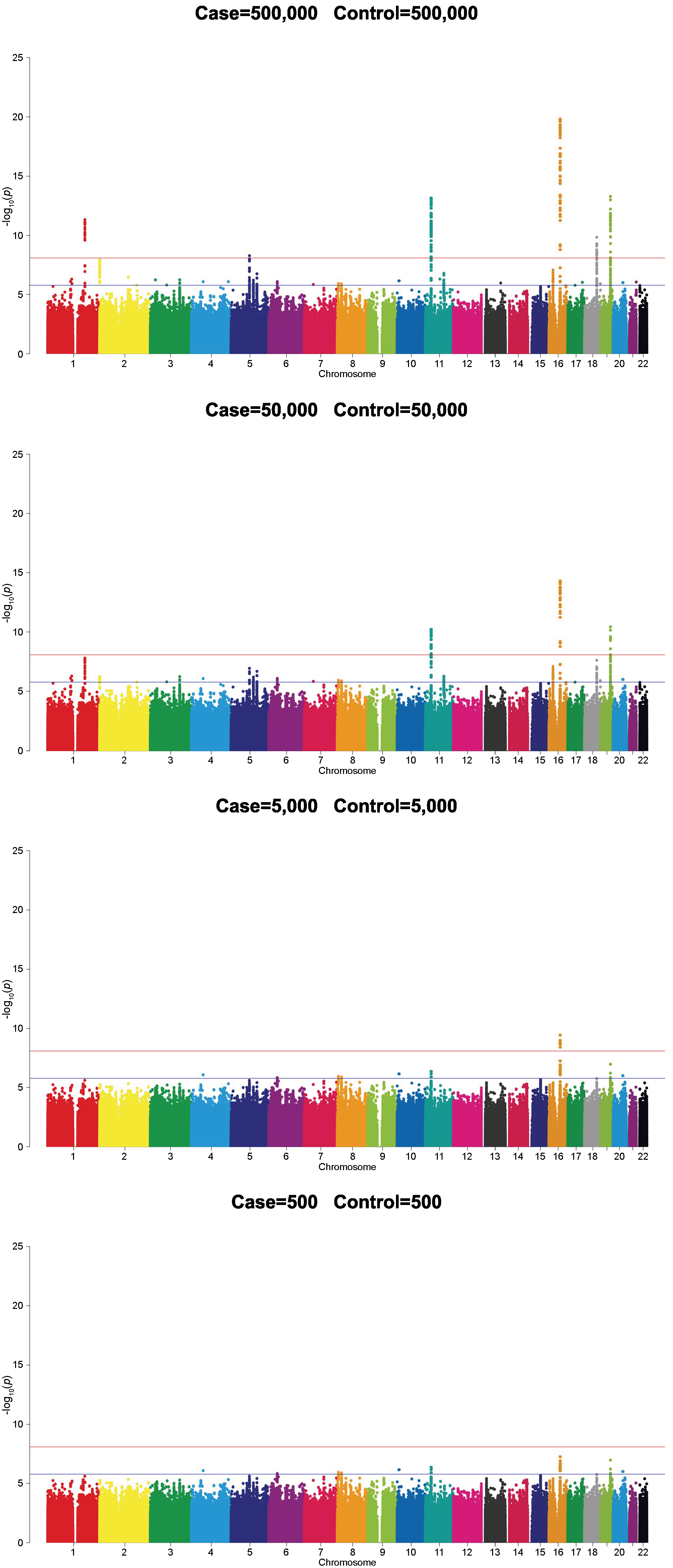

### Supplementary Figure 2. Plot of participants according to principal component 1 and 2 by principal component analysis.

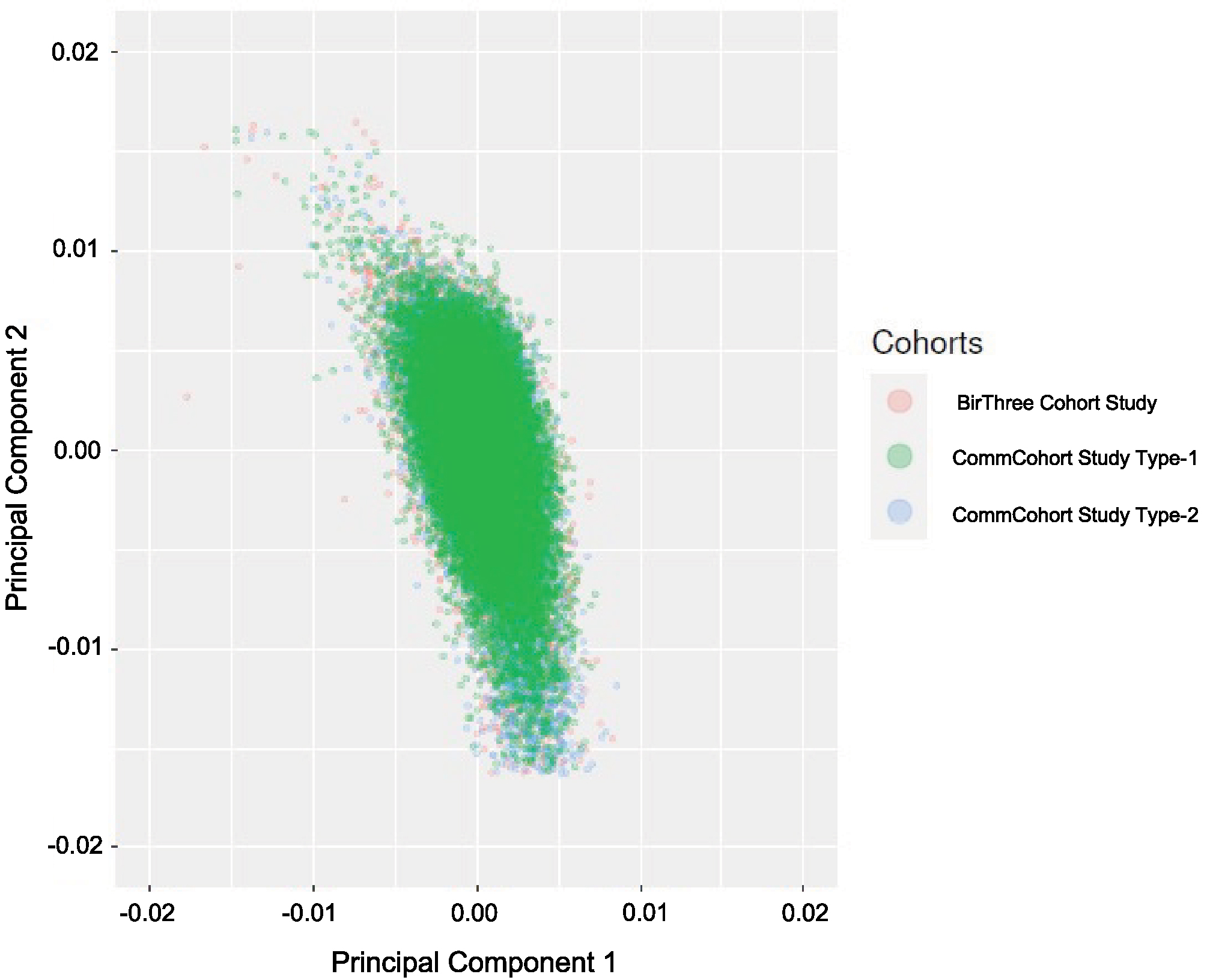

### Supplementary Figure 3. Details of the cluster-based GWAS.

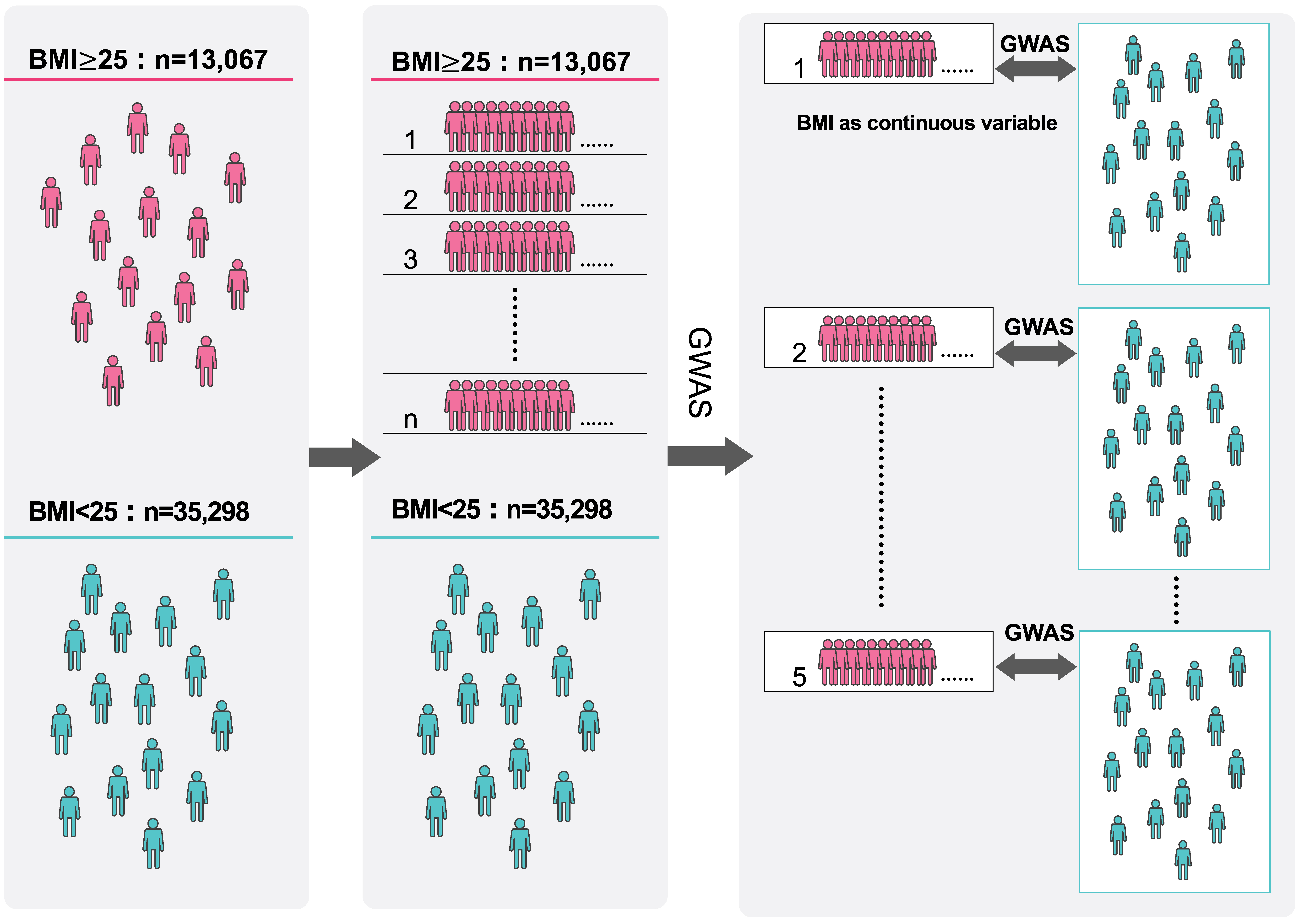

### Supplementary Figure 4. quantile-quantile plots and lambda values in main study.

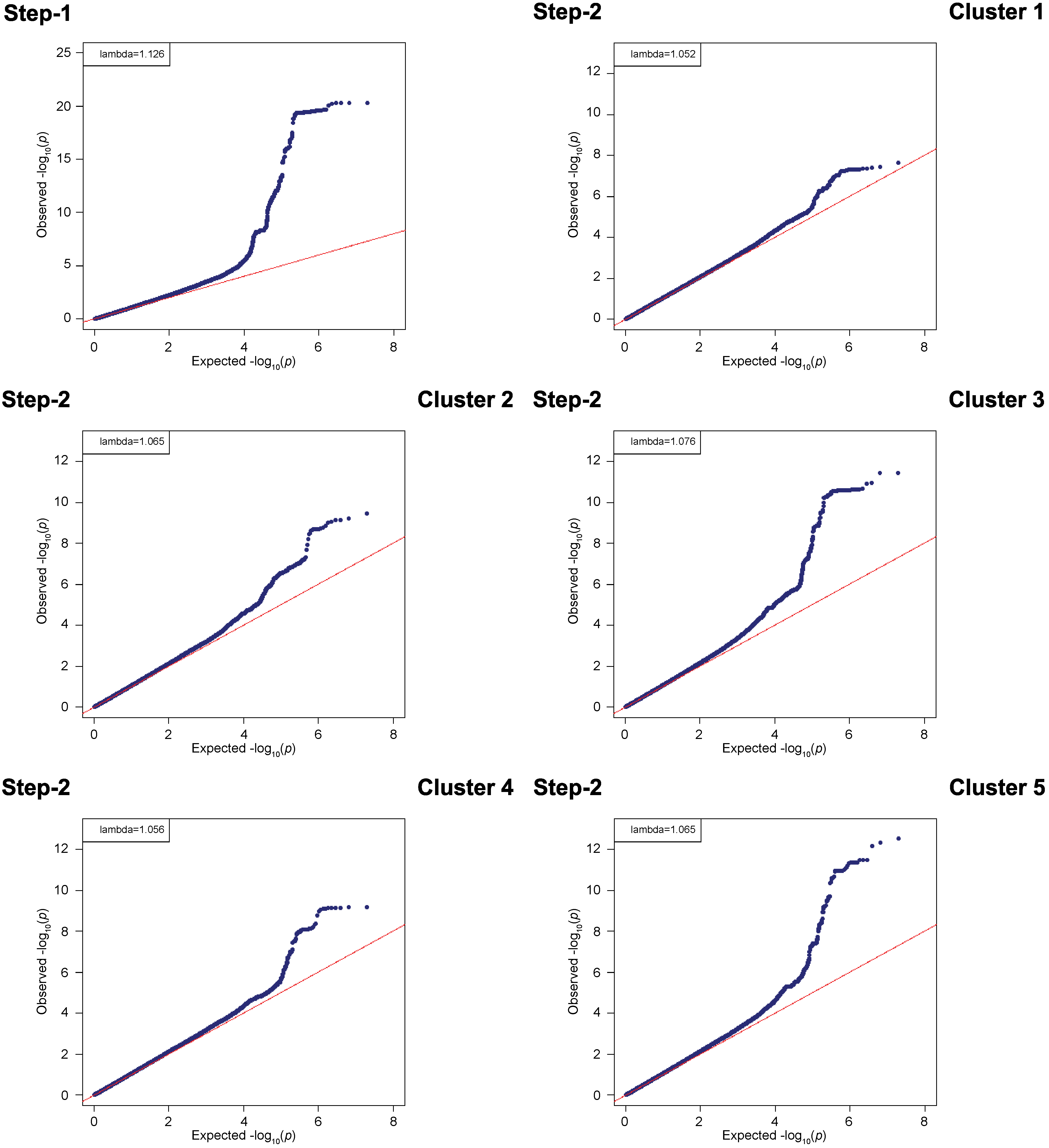

### Supplementary Figure 5. Manhattan plots (a) and corresponding quantile-quantile plots (b) of Step-1 in sub-analysis.

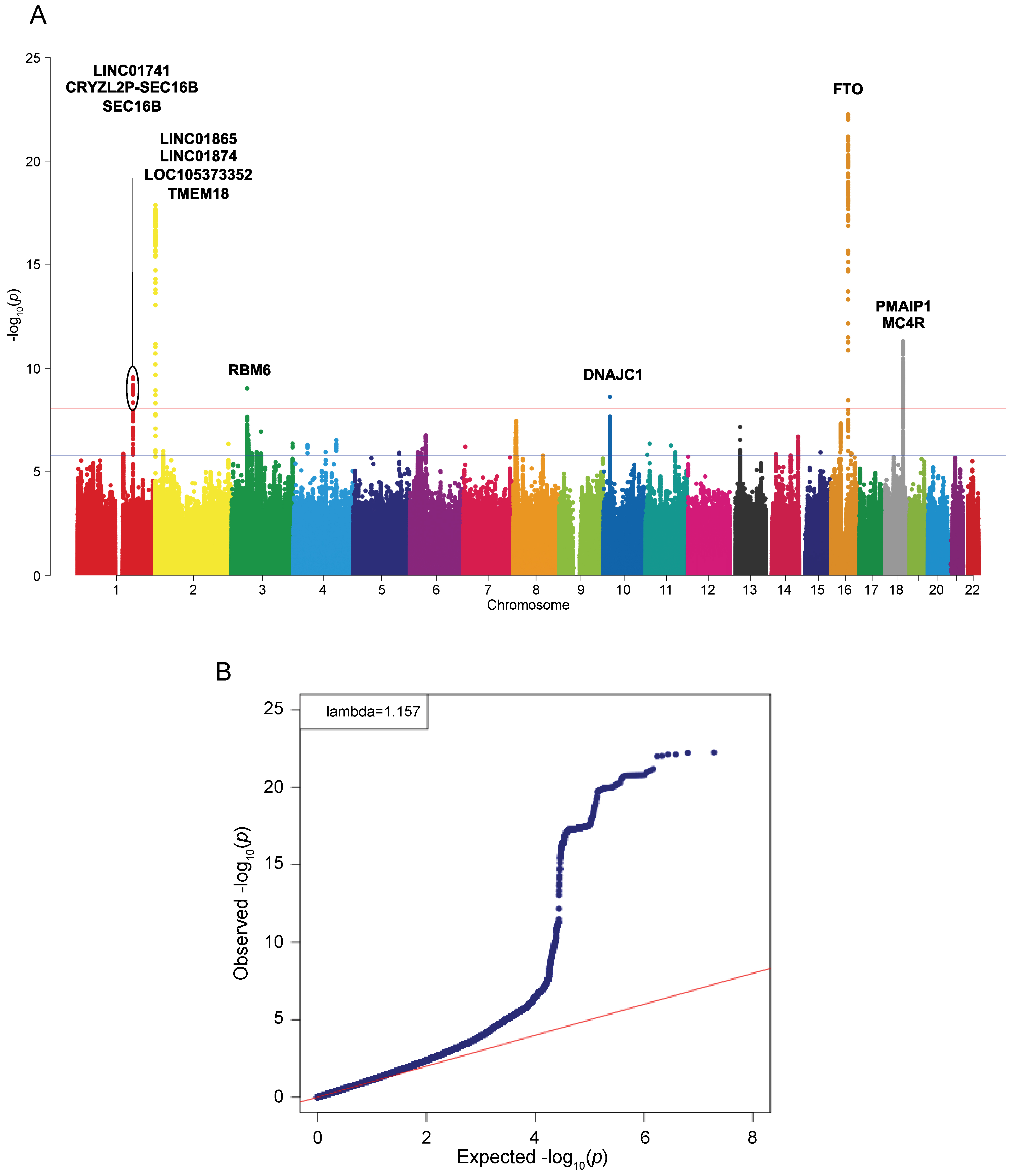

### Supplementary Figure 6. Manhattan plots (a) and corresponding quantile-quantile plots (b) of Step-2 in sub-analysis.

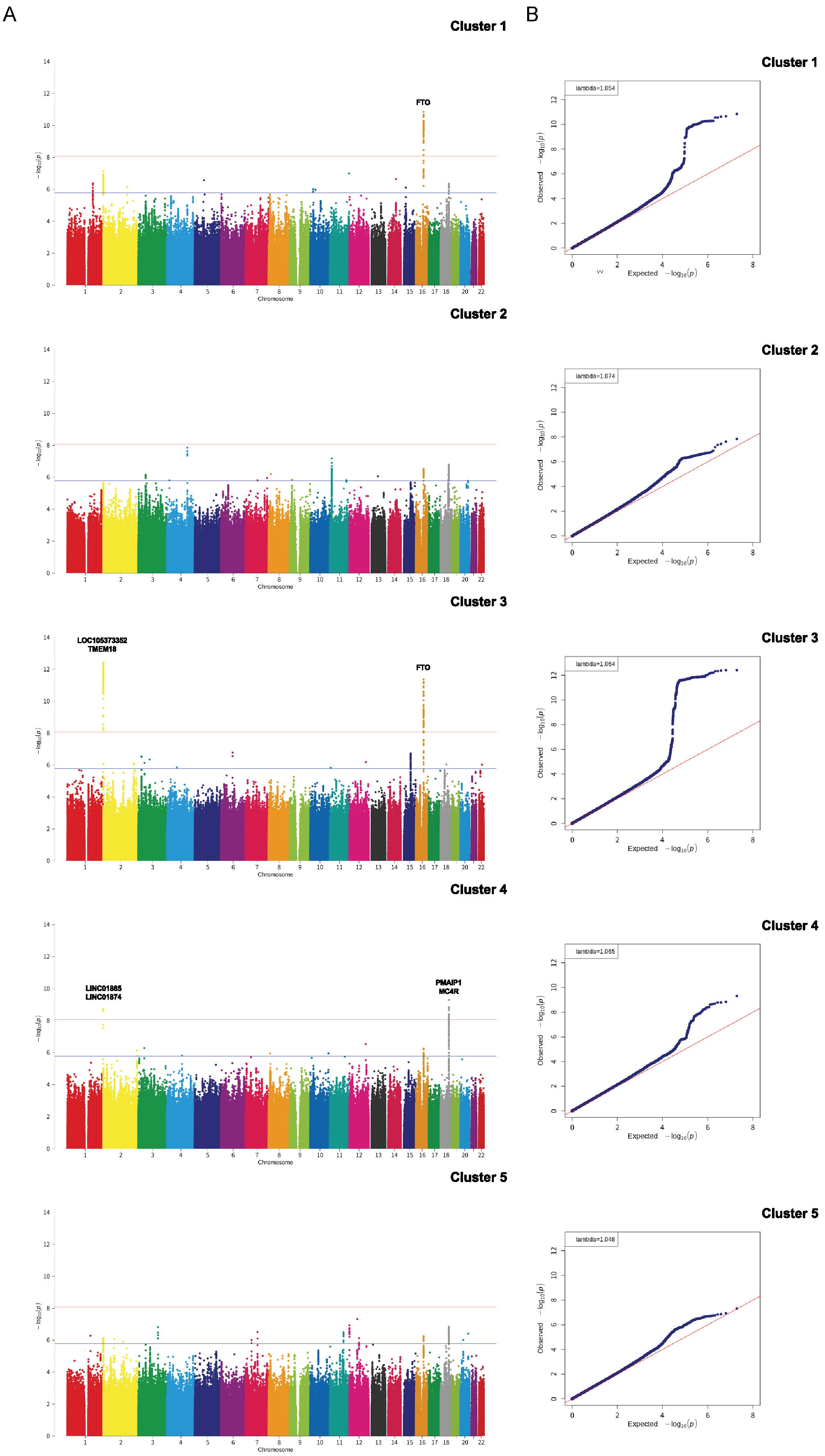
