## Supplementary documentation for "Genome wide association study based on clustering by obesity-related variables shed light on a genetic architecture of obesity in Japanese and UK population"

**Supplementary Information**

**Methods**

**Population**

Sub-analysis was based on the UK Biobank (UKB) (23–25), and all ethical regulations related to the UKB collaboration were followed. The UKB variables are managed primarily by the Category ID and corresponding Data Field IDs. In this Methods section, we describe the variables by category ID or data field ID.

The UKB is a genomic cohort of over 500,000 individuals residing in the United Kingdom (UK) between the ages of 40 and 69 years (23). Participants were recruited from 22 centers in the UK between 2006 and 2010 (23). The participants' data (*n* = 502,413) were excluded based on the following criteria: missing genetic information (*n* = 14,036), non-UK ancestry (*n* = 78,826: Data-Field 22006), pregnant at baseline (*n* = 248: Data-Field 3104), missing data on body mass index (BMI) (*n* = 1289: Data-Field 21001), BMI < 18.5 (*n* = 2,047), outliers in heterozygosity and missing rates on genetic data (*n* = 721: Data-Field 22027). Consequently, 405,246 participants were included in the study. To match the detection power of the main study and the sub-analysis, 48,365 participants were randomly selected from the 405,246 participants. For random sampling, the “sample()” function in R was used.

**Genotyping, imputation, and quality control**

The UKB participants were genotyped using the Affymetrix UK Biobank Axiom array and the Affymetrix UK BiLEVE Axiom array. Quality control, phasing, and imputation of genotyped data have been described elsewhere^2,3^. We applied the following quality controls to the post-imputation genotype dataset provided by the UKB (Category 100319): MAF < 0.01, imputation information score < 0.8. We obtained an imputed genotype dataset for 48,365 participants with 9,606,536 SNPs.

**Variables**

We selected variables for clustering from the UKB baseline survey that were similar to those in the main study. The variables included: age at recruitment (Category 21022), food intake of touchscreen questionnaire on lifestyle and personal exposures (cooked vegetables, salad/raw vegetables, fresh fruit, dried fruit, oily fish, non-oily fish, processed meat, poultry, beef, lamb/mutton, pork, cheese, bread, cereal: Category 100052), physical activity in the last year (total metabolic equivalent (MET) minutes per week for moderate activity, vigorous activity, walking: Data-Field 22027, 22028 and 22029), sleep duration (continuous variable: Data-Field 1160), smoking (never, previous, current: Data-Field 20116), alcohol drinking (never, previous, current: Data-Field 20117). The missing variables used for clustering were imputed using the k-nearest neighbor (KNN) algorithm, as in the main study (35). The KNN was implemented using the R package VIM (36) and k was set to 219.

**Clustering**

Clustering was performed in the same manner as that in the main study. Briefly, k-prototypes were used for clustering (39,40), and the number of clusters was set to 5. Continuous variables were standardized by subtracting the mean of each variable and dividing it by the standard deviation prior to clustering.

**Genome-wide association study**

The GWASs for the sub-analyses were conducted in the same manner as in the main analysis. As in Step-1, the GWAS was performed on all eligible 48,365 participants. As in Step-2, we initially clustered 32,782 of the 48,365 participants with obesity (BMI ≥ 25 kg/m^2^) using the k-prototype. Thereafter, participants in each of the five clusters of obesity and those with a BMI < 25 kg/m^2^ were combined, and a GWAS was performed according to the five clusters. To identify associations between autosomal SNPs and BMI, we used fastGWA with the GCTA software (42). The top 20 principal components calculated from the principal component analysis (Data-Field 22009), sex, and age were included as covariates. The genome-wide significance level was set to *P* < 8.33 × 10^-9^. Detected SNPs were annotated using ANNOVAR software (43). Manhattan plots and quantile-quantile plots (Q-Q plots) were generated using R (version 4.1.0).
